# Polygenic risk of obesity and BMI trajectories over 36 years: a longitudinal study of adult Finnish twins

**DOI:** 10.1101/2023.05.08.23289657

**Authors:** Bram J. Berntzen, Teemu Palviainen, Karri Silventoinen, Kirsi H. Pietiläinen, Jaakko Kaprio

## Abstract

**Objective:** We investigated 36-year body mass index (BMI) trajectories in twins whose BMI in young adulthood was below, within, or above their genetically expected BMI, with a focus on twin pairs with large intrapair BMI differences (within-pair ΔBMI ≥ 3 kg/m^2^).

**Methods:** Together, 3227 like-sexed twin pairs (34% monozygotic [MZ]) were examined at age ∼30 in 1975 and followed up in 1981, 1990, and 2011. In 1975, the observed BMI of an individual was considered either within (±2.0 kg/m^2^), below (<-2.0 kg/m^2^), or above (>+2.0 kg/m^2^) genetically expected BMI, measured by a polygenic risk score of 996,919 single nucleotide polymorphisms.

**Results:** In MZ and DZ twin pairs with large intrapair BMI differences, the co-twin with a higher observed BMI in 1975 deviated above expected BMI more frequently (∼2/3^rd^) than the co-twin with a lower BMI deviated below expectation (∼1/3^rd^). Individuals below, within, and above expectation in 1975 reached, respectively, normal weight, overweight, and obesity by 2011, with a mean BMI increase of 4.5 (95% confidence interval 4.3 to 4.8) kg/m^2^.

**Conclusion:** Categorizing BMI as below, within, or above PRS-predicted BMI helps identifying individuals who have been resistant or susceptible to weight gain. This may provide new insights into determinants and consequences of obesity.

## Introduction

Obesity prevalence has increased worldwide between 1975 and 2011 from 3.0% to 11.6% in men and from 6.6% to 15.7% in women (1). Most people living with obesity have multifactorial obesity (2), meaning it results from a complex interaction between multiple genetic loci, epigenetic factors, and environmental influences.

Twin studies have demonstrated that genetic factors explain nearly 80% of individual variation of body mass index (BMI) in young adulthood (3). Genome-wide association studies have found a large number of common genetic variants associated with BMI (4). The identified single nucleotide polymorphisms (SNPs) can be used to calculate a polygenic risk score (PRS) for BMI, which provides a personalized estimate of genetic susceptibility to obesity.

Monozygotic (MZ) co-twins have a virtually identical genetic sequence. This unique setting allows the investigation of multiple pairs of two individuals with the same PRS for BMI. Additionally, dizygotic (DZ) twin pairs share, on average, 50% of their segregating genes (i.e., PRS correlates around 0.5). When reared together, both MZ and DZ pairs are exposed to similar environmental influences from their family, neighborhood, and school, for example. Comparing MZ and DZ twin pairs helps disentangle genetic and environmental contributions to BMI. Previously, we and others have studied exceptional adult MZ and DZ twin pairs with large within-twin-pair differences in BMI (5, 6, 7, 8, 9, 10). However, these studies were cross-sectional, had various definitions for large intrapair BMI differences, and did not consider the genetic predisposition to obesity.

Large within-twin-pair BMI differences can be a transitory phenomenon as the weight of one or both siblings may fluctuate over time. Although the co-twin with a higher BMI is often considered to have gained weight relative to their sibling, this may not always be the case. In some cases, the twin with the higher BMI may have a body weight that aligns with their genetic predisposition, while the twin with a lower BMI may have a body weight below their genetically expected BMI. The latter group of co-twins either have lost weight from their natural ‘genetic level’ or have never reached their expected BMI level, suggesting they have been experiencing a protection from weight gain. Thus, earlier studies on twin pairs with large intrapair BMI differences (5, 6, 7, 8, 9, 10) have not established whether the co-twin with higher or lower BMI is the one who deviates more from their genetic predisposition.

Firstly, we will categorize individuals by whether their observed BMI falls below, within, or above their genetically expected BMI, as predicted by a whole-genome PRS. Secondly, we will give an example on how to separate twin pairs with large intrapair BMI differences into groups of twin pairs that include a co-twin resistant to weight gain versus twin pairs that include a co-twin susceptible to weight gain. Similarly, we will identify twin pairs with small intrapair BMI differences in which both co-twins have been prone to or protected against weight gain. Thirdly, we will investigate the new groups’ 36-year BMI trajectories.

## Methods

### Participants

The twin pairs in this 36-year longitudinal study were selected from the Older Finnish Twin Cohort, established in 1974 (11) and consisting of twins born before 1958 and alive in 1974 in Finland (Figure 1). The 1975 and 1981 surveys targeted all twins in the cohort, while the 1990 survey was restricted to twins born 1930–1957. Genotype data were collected mainly from the late 1990s onwards. Of the twins participating in 1975, 3,227 complete same-sex twin pairs had genotype data (34% MZ). The 2011 data collection targeted twins born 1945–1957, with 943 of them (44% MZ) having genotype data. Data collection was approved by the ethics committee of the Hjelt Institute, University of Helsinki and the ethics committee of the Helsinki and Uusimaa Hospital District, Finland and the research was conducted in accordance with the principles of the Declaration of Helsinki. All participants gave informed consent.

**Figure 1:**
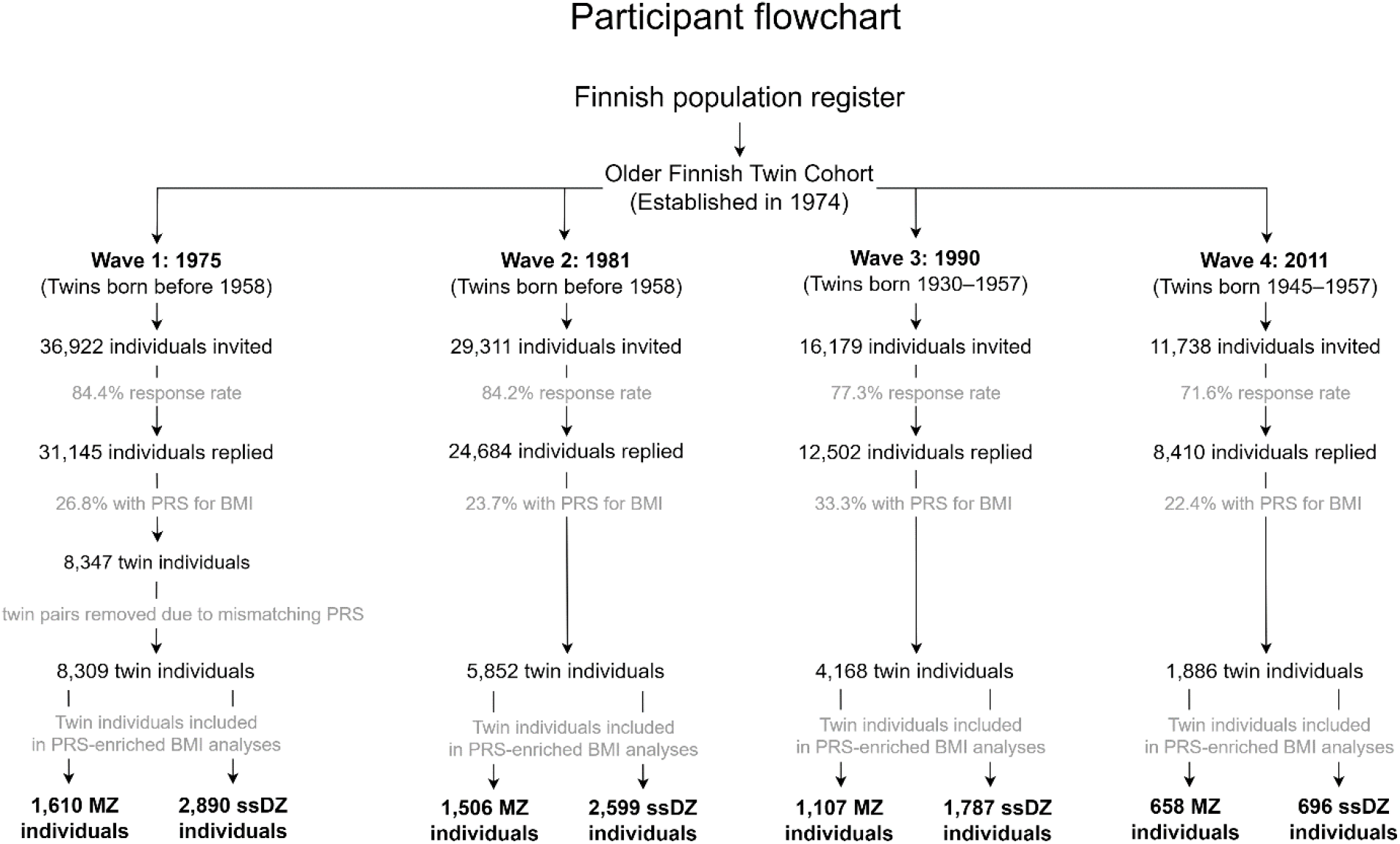
PRS = polygenic risk score, BMI = body mass index, MZ = monozygotic, ssDZ = same-sex dizygotic.

### Twin characteristics

Zygosity was confirmed through genotyping information derived from blood samples. We removed 19 MZ pairs (taken away from the original MZ sample size, n = 1105) with low pi-hat values for relatedness despite being labeled as MZ pair, mostly due to sample mix-up. Personal characteristics were self-reported through a questionnaire. BMI based on self-reported weight and height is validated in this cohort and found to be highly reliable with a correlation of 0.90 in women and 0.89 in men (12). BMI was categorized as underweight (<18.5), normal weight (18.5 to <25), overweight (25 to <30), obesity (30+) (13).

### Polygenic risk score calculation

The PRS for BMI was based on 996,919 common SNPs (minor allele frequency > 5% in Europeans), calculated using HapMap3 SNPs, with 27,284 FINRISK individuals as a linkage disequilibrium reference panel (14, 15). FINRISK is another Finnish cohort without sample overlap with our twin cohort. LDpred and PLINK were used to calculate the PRS. We created standardized values (z-scores) for PRS to simplify interpretation.

### Statistical analyses

Stata/MP 17.0 (College Station, TX: StataCorp LLC) was used for statistical analyses. We performed a linear regression model to estimate how well the PRS for BMI predicted BMI with age and sex as covariates. From the regression model, we predicted BMI values and calculated how much the observed BMI differed from the predicted BMI. Predicted BMI will be referred to as “expected BMI”. Subsequently, the size and direction of the differences between observed and expected BMI were used to develop novel groups of twin pairs discordant for PRS-enriched BMI (two groups: one co-twin 1. above or 2. below expected BMI, with their counterpart’s BMI within expectation) and twin pairs concordant for PRS-informed BMI (three groups: both co-twins 1. within, 2. below, or 3. above expected BMI), as elaborated upon in the results. We used independent t-tests to compare differences between MZ and DZ pairs.

Repeated measures mixed-effects linear regressions with unstructured covariance and maximum likelihood estimation were performed to compare the observed BMI trajectories between co-twins. This method used all available datapoints regardless of missing values at one or more visits. We performed one model without specifications for the unadjusted values in BMI trajectories, and one adjusted model that identified repeated measures of individuals and clustering within twin pairs. Only the adjusted values were reported in the main text. We used the “contrast” command to compare within-pair BMI differences at each timepoint and to estimate whether a significant change in BMI occurred between baseline and final follow-up in each group of co-twins. We performed post-hoc tests of partial interaction between intrapair BMI differences and adjacent year of visit (1975 vs 1981, 1981 vs 1990, and 1990 vs 2011). Time points were coded as zero for 1975, six for 1981, fifteen for 1990, and thirty-six for 2011.

## Results

### Demographics

The 6,454 twin individuals from the complete twin pairs in 1975 were in their thirties, on average, and in 2011 the remaining 1,886 individuals were around age sixty (Table 1). The 36-year follow-up showed a lower average increase in age (60.5 – 33.4 = 27.1 y) due to a higher drop-out rate among older individuals, which resulted from study design exclusions and mortality. The study had a balanced sex representation, with over 40% of participants being men at all surveys. In 1975, 1981, and 1990, normal weight was the norm, but by 2011 overweight surpassed normal weight. Over the 36-year period, all individuals gained, on average, 3.2 kg/m^2^. The BMI categories were similar to those measured by NCD-RisC (and age-standardized in adults over age 20) in the same years in Finland (Table S1), except the obesity prevalence was notably lower in most years in our study sample. In 1975, about 25% of the twin pairs had large intrapair BMI differences (≥ 3 kg/m^2^; of which 18% were MZ and 82% DZ). This proportion increased to nearly 50% in 2011 (of which 33% were MZ and 67% DZ). A total of 405 MZ pairs (37%) and 1230 DZ pairs (57%) had large within-pair differences in BMI at some point during the study.

**Table 1:** Characteristics of twin individuals from complete twin pairs per timepoint

|  | 1975<br>n = 6454 | 1981<br>n = 5852 | 1990<br>n = 4168 | 2011<br>n = 1886 |
| --- | --- | --- | --- | --- |
| Age in 1975, years | 33.4 (9.4) | 39.6 (9.5) | 47.4 (7.7) | 60.5 (3.7) |
| Women/men (female %) | 3,584/2,870 (56) | 3,308/2,544 (57) | 2,396/1,772 (57) | 1,088/798 (58) |
| Zygoty, MZ freq. (%) | 2,172 (34) | 2,018 (34) | 1,500 (36) | 824 (44) |
| Observed BMI, kg/m<sup>2</sup> | 23.2 (3.2) | 23.9 (3.4) | 25.0 (3.9) | 26.4 (4.4) |
| Underweight, freq. (%) | 267 (4) | 145 (2) | 69 (2) | 18 (1) |
| Normal weight, freq. (%) | 4,535 (70) | 3,785 (65) | 2,227 (53) | 746 (40) |
| Overweight, freq. (%) | 1,441 (22) | 1,639 (28) | 1,490 (36) | 786 (42) |
| Obesity, freq. (%) | 211 (3) | 283 (5) | 382 (9) | 336 (18) |
| Intrapair BMI difference $\geq 3$ kg/m<sup>2</sup>, freq. (%) | 1,634 (25) | 1,618 (28) | 1,432 (34) | 836 (44) |

Values are mean (SD) or frequency (percentage). n = number of individuals, MZ = monozygotic, BMI = body mass index. BMI categories (kg/m<sup>2</sup>): underweight (<18.5), normal weight (18.5 to <25), overweight (25 to <30), obesity (30+).

### BMI trajectories by top and bottom PRS deciles

BMI differences between top and bottom PRS deciles were substantial at all timepoints (p<0.001; Figure 2 and Table S2a&b; mean BMI and participants per PRS decile are shown in Table S3a–c). The post hoc test for partial interaction provided evidence (p < 0.01) that persons in the top 10% of PRS showed a larger increase in BMI over 36 years (5.4 kg/m^2^: from 24.6 to 30.0 kg/m^2^) than did persons in the lowest 10% (3.6 kg/m^2^: 22.1 to 25.7 kg/m^2^).

**Figure 2:**
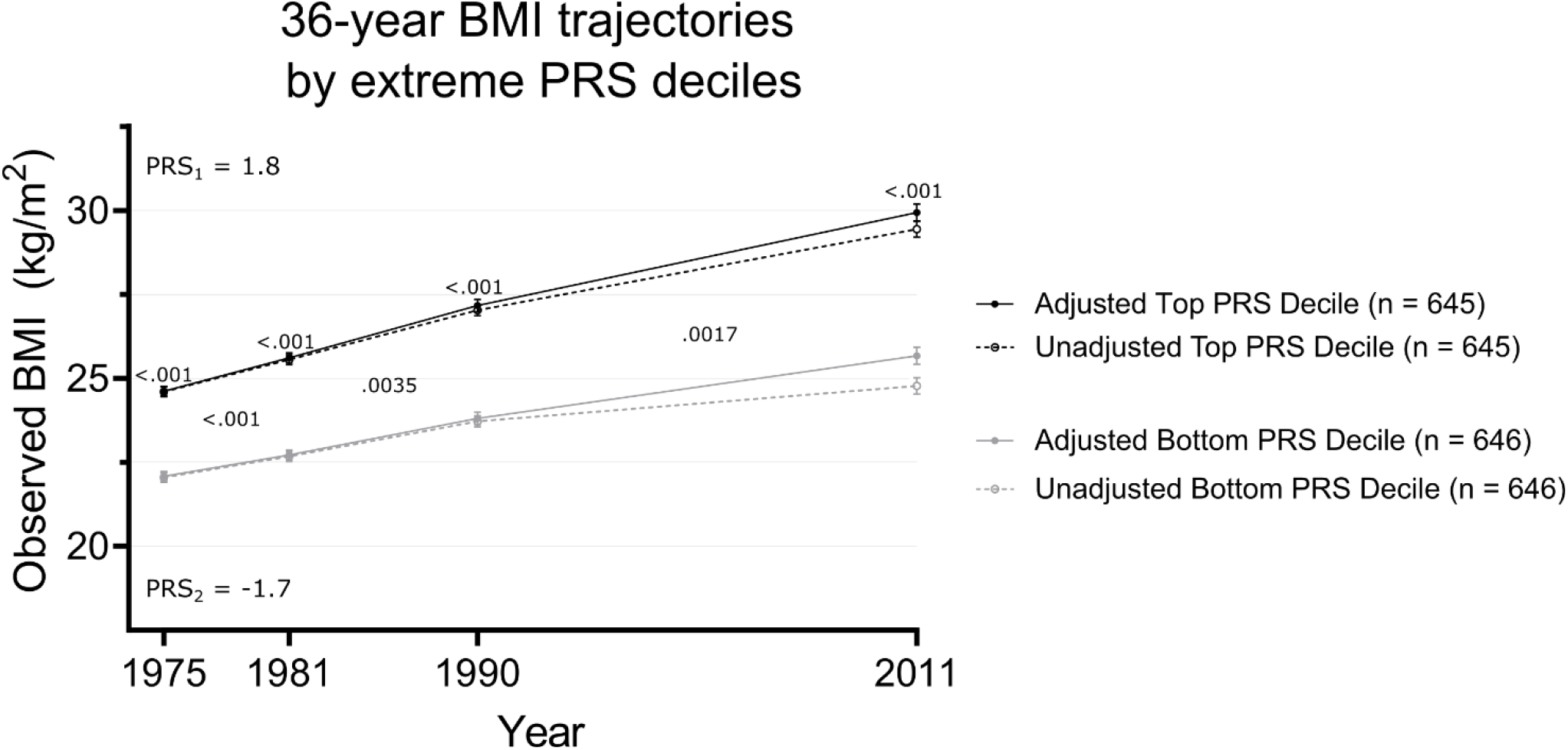
Unadjusted and adjusted values of observed body mass index (BMI) trajectories in top and bottom polygenic risk score (PRS) deciles of all twin participants. PRS_1_ = mean PRS of individuals in the top PRS decile, PRS_2_ = mean PRS of individuals in the bottom PRS decile. Adjusted p-values below 0.10 are displayed in the graph, derived from contrast tests and post-hoc tests of partial interaction from the repeated measures mixed-effects linear regressions.

### Expected BMI in 1975 based on PRS and observed BMI in 1975

In 1975, the PRS independently explained about 4.9% of the variation in BMI (Table S4a). However, when corrected for age and sex, PRS predicted 29.3% of the BMI variation (Table S4b). With this corrected model, we predicted the genetically expected BMI. On average, the observed BMI differed 2.0 kg/m^2^ from the expected BMI (Figure S1A&B). An individual’s BMI in 1975 was categorized as below expectation (observed BMI more than 2.0 kg/m^2^ below the expected BMI), within expectation (observed BMI within 2.0 kg/m^2^ from the expected BMI in either direction), or above expectation (observed BMI more than 2.0 kg/m^2^ above the expected BMI).

### Developing twin pair groups with PRS-based discordance and concordance for BMI

We categorized twin pairs based on observed BMI and expected BMI calculated by PRS. Firstly, we grouped twin pairs as having either large (ΔBMI ≥ 3 kg/m^2^) or small (ΔBMI < 3 kg/m^2^) observed BMI differences within pairs. Additionally, we evaluated how both co-twins’ observed BMI related to their genetically expected BMI in 1975 (Example in Figure 3; scatterplot of all datapoints by zygosity in Figure S2). We refer to these new groups as twin pairs discordant and concordant for PRS-enriched BMI. Two groups of twin pairs discordant for PRS-enriched BMI were created: a) one co-twin with observed BMI within expected BMI range, the other co-twin with observed BMI above expected BMI, b) one co-twin within BMI expectation, the other co-twin below expectation. Three groups of twin pairs concordant for PRS-enriched BMI were developed, with both co-twins’ observed BMI: a) within, b) below, or c) above expected BMI. All pairs who failed to fulfill these group requirements were excluded. In comparison with large or small intrapair differences based on observed BMI alone, this new selection captured 109/150 (73%) MZ and 468/667 (70%) DZ pairs with large intrapair BMI differences and 696/936 (74%) MZ and 977/1474 (66%) DZ pairs with small intrapair BMI differences, which left a total of 4,500 individuals in 1975 for further investigation. All groups included individuals over a wide range of PRS scores (Figure S2).

**Figure 3:**
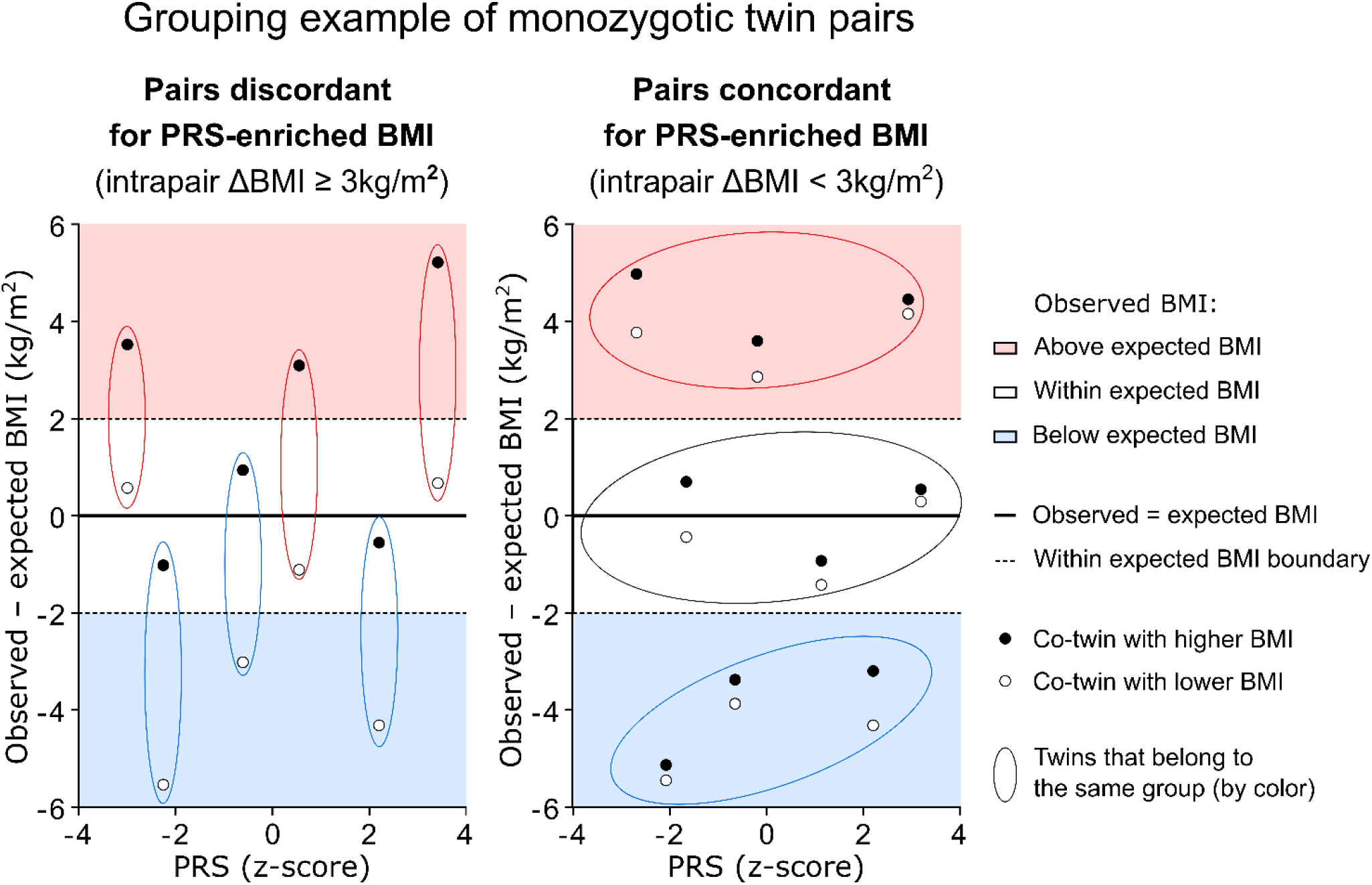
Example of the development of twin pair groups discordant and concordant for PRS-enriched BMI based on the deviation between observed body mass index (BMI) and genetically expected BMI, plotted against the polygenic risk score (PRS).

### Characteristics of twin pair groups discordant and concordant for PRS-enriched BMI

In 1975, the observed BMI of 2,845 individuals was in line with genetically expected BMI (65%), 811 deviated below (18%), and 748 (17%) deviated above BMI expectation. In the twin pairs discordant for PRS-enriched BMI, it was more common that the co-twin with higher observed BMI (1975) was the co-twin above expected BMI in both MZ (67%) and DZ (62%) twin pairs. In the remaining 33% and 38%, the co-twins with lower observed BMI were the exceptional co-twins, deviating below their BMI expectation. In most twin pairs concordant for PRS-enriched BMI, both co-twins of the pair followed their genetic predisposition to BMI in MZ (67%) and DZ (72%) pairs. The mean PRS and expected BMI were similar between newly created twin groups, and the proportion of female pairs ranged from 52–64% between groups (group details are in Tables S4–S9).

Repeated measures linear mixed models showed that observed BMI increased over time in all PRS-enriched BMI groups (p<0.001). The average [95% confidence interval, 95% CI] observed BMI increase in the 4,500 individuals from all groups combined was 4.5 [4.3, 4.8] kg/m^2^ (p<0.001). The mean observed BMI of all individuals above BMI expectation in 1975 (BMI = 28.0 kg/m^2^) passed the obesity threshold by 2011 (mean BMI = 32.5 kg/m^2^), whereas those within the range of BMI expectation in 1975 (BMI = 23.0 kg/m^2^) had on average overweight in 2011 (BMI = 27.3 kg/m^2^). The individuals whose BMI was below their genetic predisposition in 1975 (BMI = 19.6 kg/m^2^) had, on average, normal weight in 2011 (BMI = 24.1 kg/m^2^).

### BMI trajectories in twin pairs discordant for PRS-enriched BMI

We performed repeated measures linear mixed models to compare the BMI trajectories between co-twins of twin pairs discordant for PRS-enriched BMI (number of twin pairs and sex proportion by timepoint in Table S5). The BMI increases over time in the co-twin groups from twin pairs discordant for PRS-enriched BMI ranged from 3.7 to 5.5 kg/m^2^ in MZ twins and 4.2 to 5.5 kg/m^2^ in DZ twins (p<0.001 in all groups). In both MZ and DZ pairs, the co-twins below expectation stayed in the normal weight BMI category over time (except DZ co-twins reached overweight), the co-twins within expectation went from normal weight to overweight, and the co-twins above expectation went from overweight to obesity (Figure 4 and Table S10a&b–S11a&b).

**Figure 4:**
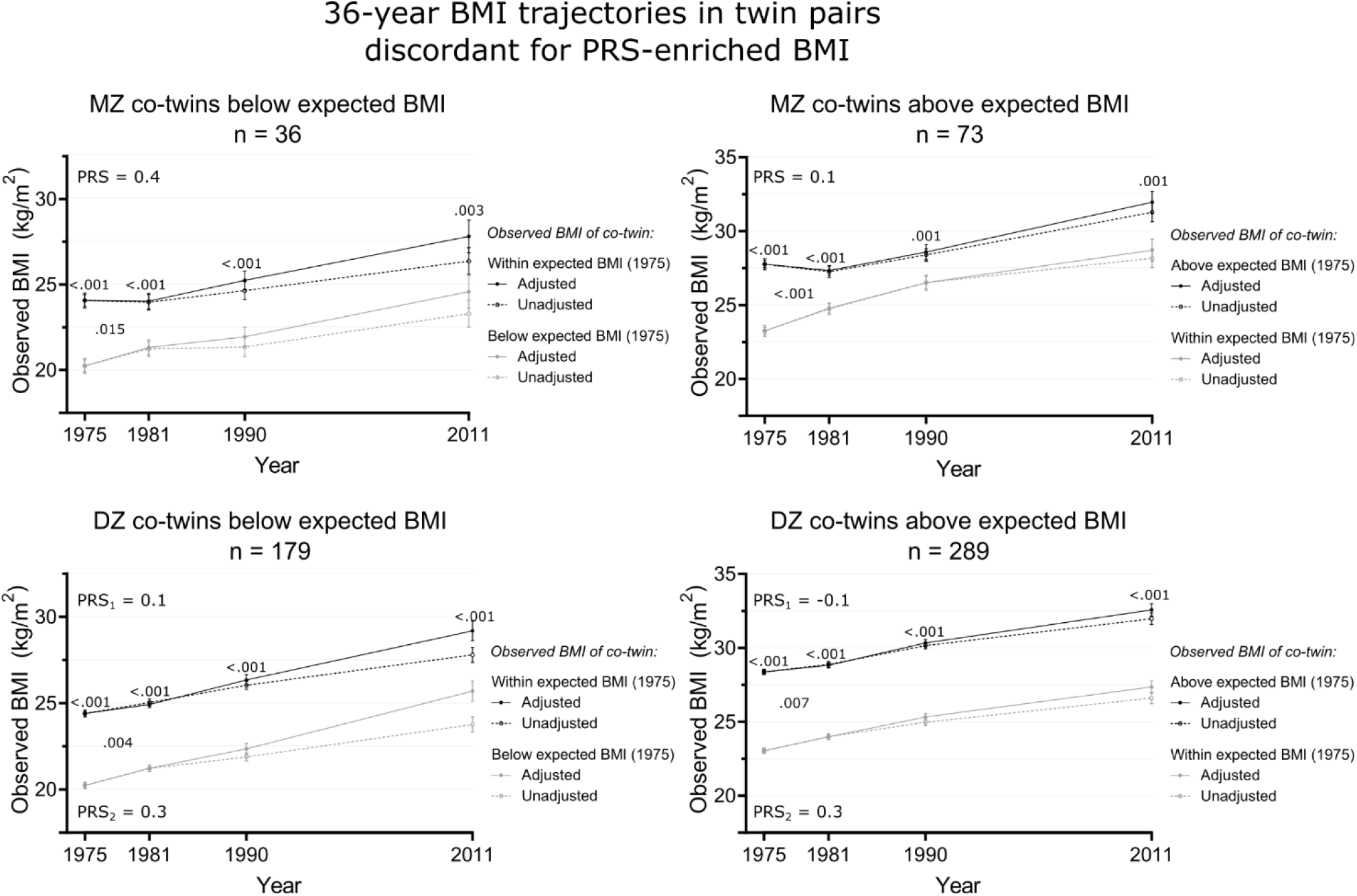
Unadjusted and adjusted values of observed body mass index (BMI) over time in monozygotic (MZ) and dizygotic (DZ) twin pairs discordant for PRS-enriched BMI (large within-twin-pair BMI differences [≥3 kg/m^2^] with an additional consideration of observed and expected BMI). Adjusted p-values on top of each timepoint reflect contrast tests of within-pair BMI differences in that year, and adjusted p-values between two timepoints reflect post-hoc tests of partial interaction effects between the BMI trajectories of the co-twins during the adjacent years. n = number of pairs, PRS = mean polygenic risk score in both MZ co-twins in a pair. PRS_1_ = mean polygenic risk score of the DZ co-twins with higher BMI in 1975, PRS_2_ = mean polygenic risk score of DZ co-twins with lower BMI in 1975. Adjusted p-values below 0.10 are displayed in the graph, derived from contrast tests and post-hoc tests of partial interaction from the repeated measures mixed-effects linear regressions.

The MZ and DZ twin pairs discordant for PRS-enriched BMI in 1975 maintained substantial intrapair BMI differences at all timepoints (MZ ΔBMI range = 2.1–4.5 kg/m^2^, p < 0.005, DZ ΔBMI range = 3.8–5.4 kg/m^2^, p < 0.001; Figure 4, Table S10a&b and S11a&b). The intrapair BMI differences were larger in DZ than MZ twin pairs discordant for PRS-enriched BMI at all timepoints (independent t-test p < 0.005), except in the pairs with co-twins below expectation in 1990 (p = 0.23) and 2011 (p = 0.18; BMI contrast values in Table S10a&b vs S11a&b). The post hoc test for partial interaction showed that the intrapair BMI differences diminished between 1975 and 1981 in all groups (Figure 4 and Table S12a&b), which appeared more pronounced in MZ than DZ twin pairs. In twin pairs containing co-twins with an observed BMI above expectation, the intrapair BMI difference decreased in MZ pairs by 2.0 kg/m^2^ (p < 0.001) and in DZ pairs by 0.5 kg/m^2^ (p = 0.004). In twin pairs with co-twins having an observed BMI below expectation, the BMI difference diminished in MZ pairs by 1.1 kg/m^2^ (p = 0.014) and in DZ pairs by 0.5 kg/m^2^ (p = 0.004).

### BMI trajectories in twin pairs concordant for PRS-enriched BMI

In twin pairs concordant for PRS-enriched BMI (number of twins and sex proportion by timepoint in Table S6), the BMI increases over time in the co-twin groups ranged from 4.0 to 4.4 kg/m^2^ in MZ twins and 4.1 to 4.9 kg/m^2^ in DZ twins (p<0.001 in all groups). Overall, in most cases, the twin pairs below expectation stayed within the normal weight category, the twin pairs within expectation went from normal weight to overweight, and the twin pairs above expectation went from overweight to obesity (Figure 5, Table S13a&b, and Table S14a&b).

**Figure 5:**
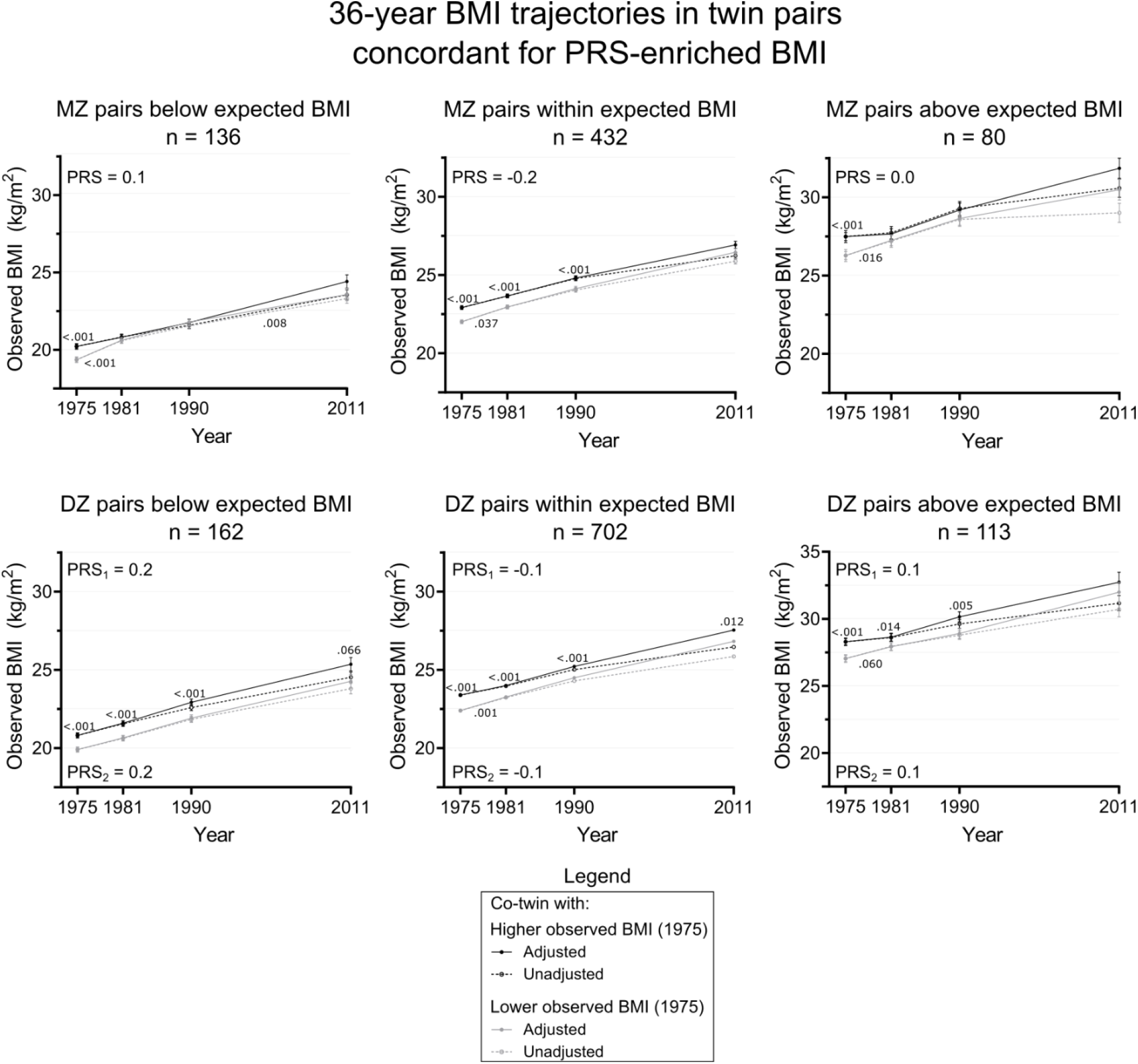
Unadjusted and adjusted values of observed body mass index (BMI) over time in monozygotic (MZ) and dizygotic (DZ) twin pairs concordant for PRS-enriched BMI (small within-twin-pair BMI differences [<3 kg/m^2^] with an additional consideration of observed and expected BMI). Adjusted p-values on top of each timepoint reflect contrast tests of within-pair BMI differences in that year, and adjusted p-values between two timepoints reflect post-hoc tests of partial interaction effects between the BMI trajectories of the co-twins during the adjacent years. n = number of pairs, PRS = mean polygenic risk score in both MZ co-twins in a pair. PRS_1_ = mean polygenic risk score of the DZ co-twins with higher BMI in 1975, PRS_2_ = mean polygenic risk score of DZ co-twins with lower BMI in 1975. Adjusted p-values below 0.10 are displayed in the graph, derived from contrast tests and post-hoc tests of partial interaction from the repeated measures mixed-effects linear regressions.

Contrast tests revealed intrapair BMI differences in 1975 in all MZ twin pair groups and in 1981 and 1990 in the MZ pairs within expected BMI. Intrapair BMI differences in DZ pairs were present at all timepoints, except in 2011 for the groups below and above expected BMI. Intrapair differences were similar between MZ and DZ twin pairs concordant for PRS-enriched BMI (BMI contrasts in Table S13a&b vs Table S14a&b), except in pairs with both co-twins below expectation in 1981 (independent t-test p < 0.001) and 1990 (p = 0.0021), when the intrapair BMI differences were ∼0.7 kg/m^2^ larger within DZ pairs. Among all types of MZ twin pairs concordant for PRS-enriched BMI, the within-pair BMI difference decreased between 1975 and 1981 (pairs below: 0.7 kg/m^2^, p<0.001; within: 0.2 kg/m^2^, p = 0.037; above: 0.8 kg/m^2^, p = 0.016; Figure 5 and Table S15a&b). Among the DZ pairs within expected BMI, the intrapair BMI differences decreased by 0.3 kg/m^2^ (p = 0.001) between 1975 and 1981 (Figure 5 and Table S15a&b), whereas no changes were seen for other DZ pairs.

## Discussion

To the best of our knowledge, this study is the first of its kind to determine in a pair of twins with large intrapair BMI differences whom of the co-twins had acquired a BMI that deviated from their genetically predetermined BMI. Individuals’ observed BMIs were labeled as being below, within, or above genetically expected BMI in 1975, based on which we defined groups of twin pairs discordant or concordant for PRS-enriched BMI. We then traced their BMI over 36 years. In pairs discordant for PRS-enriched BMI, one co-twin followed their genetic predisposition, and the other deviated either below or above expectation. While most discordant pairs contained one co-twin with BMI above genetic expectation, still about one-third of the pairs included a co-twin with a BMI far below biological predisposition. In pairs concordant for PRS-enriched BMI, most twins’ BMI (∼70%) was in line with their genetic predisposition. BMI increased similarly with time in all groups, thus the BMI during young adulthood at the first visit appeared to be the main determinant of BMI at the 36-year follow-up visit. The group of individuals with a BMI below genetic expectation maintained, on average, a normal weight over 36 years, whereas groups consistent with or above expectation reached overweight and obesity, respectively.

The observed BMI of the divergent co-twin in MZ and DZ twin pairs discordant for PRS-enriched BMI deviated more commonly above expected BMI (∼2/3^rd^) than below (∼1/3^rd^) in 1975. Thus, in MZ twins who share an identical genetic sequence, discordance for BMI in young adulthood more likely arose from environmental influences that stimulated weight gain in one of the co-twins rather than protecting from weight gain or promoting weight loss. By grouping individuals’ BMI by their biological predisposition to BMI, we can distinguish people who have been susceptible to weight gain or not, regardless of their genetic risk. These two different types of twin pairs discordant for PRS-enriched BMI could form the basis of further studies into etiological determinants of body weight and weight change, regardless of genetic influences.

BMI increased on average 4.5 kg/m^2^ between 1975 and 2011 in all 4,500 individuals from the PRS-enriched BMI groups. BMI is known to increase with aging from early (18–25 y) to middle (65 y) adulthood with about 3.0–3.5 kg/m^2^ in men and 4.0–5.7 kg/m^2^ in women, all born before 1958, as in our study (18, 19). The increasing obesogenic environment in Finland may also contribute, which can be derived from the increasing age-standardized adult obesity rates in Finland between 1975 and 2011 (Table S1, (1)). This was further exemplified by a Norwegian study showing that BMI throughout adulthood was higher in individuals born after 1970 compared to before, regardless of genetic predisposition to obesity (16).

Most twin individuals (64%) approximated their genetic predisposition to BMI during young adulthood as expected from the high heritability of BMI (3). The genetic drive towards BMI was supported by the MZ-DZ-comparisons in our study. Firstly, the DZ pairs more frequently had intrapair BMI differences over 3 kg/m^2^ at some point in time than MZ pairs (57% vs 37%). Secondly, the intrapair BMI differences in MZ twin pairs discordant for PRS-enriched BMI were generally smaller than those of DZ pairs. Thirdly, the intrapair differences in twin pairs discordant for PRS-enriched BMI seemed to decrease more strongly in MZ than DZ pairs between 1975 and 1981, displaying a trend toward a more similar BMI in MZ than DZ pairs. Furthermore, in agreement with studies linking a higher genetic obesity susceptibility to a steeper BMI trajectory (16, 17), we found that individuals in the lowest genetic risk decile increased only 3.6 kg/m^2^ over 36 years versus 5.4 kg/m^2^ in the highest decile.

While twin individuals in the highest PRS decile showed a steep upward BMI trajectory and attained a mean BMI of 30.0 kg/m^2^ in 2011, a similar number of individuals deviated above their genetically expected BMI and crossed the obesity threshold in 2011 as well (32.5 kg/m^2^). On the other end, twins in the bottom PRS decile had a modest increase in BMI and acquired overweight (25.7 kg/m^2^), but twins with an observed BMI below expectation maintained a normal overweight up until 2011 (24.1 kg/m^2^). Genetic risk was similar between twins with observed BMI below and above expected BMI, confirming that genetic risk is probabilistic, not deterministic, and highlighting the important contribution of the environment to development and trajectories of weight gain and loss. Future studies should investigate the determinants and health consequences of low, middle, and high BMI trajectories consistent with genetic risk, in comparison to trajectories below or above biological predisposition.

This study has both strengths and limitations. The research design consisted of a large prospective cohort of twin pairs, who were examined on four occasions over the course of 36 years, with DNA samples collected from 3227 complete twin pairs, which were used to calculate a recent genome-wide PRS. However, the PRS still needs improvement as an independent predictor, as in its current form it accounted for a relatively small proportion of the variation in BMI (∼5%). Then again, after inclusion of age and sex in the model, the PRS prediction of BMI variation was substantial (∼29%), similar to the SNP-based heritability via whole-genome sequencing of 0.30 (20). The genetically expected BMI in our study is not a fixed value based solely on DNA but depends on the environment and study sample at a certain time. We further acknowledge that BMI is an imperfect determinant of obesity in an individual, but we used it as a crude indication of adiposity differences at the group level.

## Conclusion

Via the categorization of BMI as below, within, or above genetically expected BMI, we identified twins who presumably have been protected from or prone to weight gain up until young adulthood. In about two-third of twin pairs discordant for PRS-enriched BMI, the co-twin with higher BMI diverged above their genetically expected BMI, whereas in one-third of the pairs, the co-twin with lower BMI was below genetic expectation. Grouping individuals by their observed BMI in relation to their genetically expected BMI opens avenues for research into weight gain susceptibility. After categorization, BMI steadily increased ∼4.5 kg/m^2^ with ageing in most groups between 1975 and 2011, regardless of genetic risk or BMI. Due to the steady BMI increase, the BMI level in 1975 mainly determined the BMI level in 2011. Twins below BMI expectation maintained a normal weight in over time, in contrast to twins within expectation who went from normal weight to overweight, and twins above expectation who went from overweight to obesity. The determinants and health implications of regular BMI trajectories versus PRS-informed BMI trajectories require further investigation.

## Data Availability

The twin dataset used in the current study will be located in the Biobank of the Finnish Institute for Health and Welfare, Finland. All the biobanked data are publicly available for use by qualified researchers following a standardized application procedure (https://thl.fi/en/web/thl-biobank/for-researchers).

## Acknowledgements

We would like to thank the study participants for their generous, long-term, and invaluable contributions to our research.

