## Supplemental material for "Polygenic risk of obesity and BMI trajectories over 36 years: a longitudinal study of adult Finnish twins"

### Online Supporting Information

Table S1: Prevalence of BMI categories in Finland in 1975, 1981, 1990, and 2011 according to NCD-RisC statistics, age-standardized among adults aged 20+ (1)

| | BMI category prevalence in Finland | | | |
|---|---|---|---|---|
| | 1975 | 1981 | 1990 | 2011 |
| Underweight, % | 2 | 2 | 1 | 1 |
| Healthy weight, % | 62 | 58 | 51 | 41 |
| Overweight, % | 29 | 32 | 35 | 37 |
| Obesity, % | 7 | 9 | 13 | 21 |
| Mean BMI, $kg/m^2$ | 24.5 | 24.8 | 25.3 | 26.1 |

Values are percentages or mean. BMI = body mass index. BMI categories ($kg/m^2$): underweight (<18.5), normal weight (18.5 to <25), overweight (25 to <30), obesity (30+).

Table S2a: Unadjusted values of characteristics of twin individuals in bottom and top PRS deciles

| Comparison of characteristics between twins in top and bottom PRS decile | | | | | | | | | | | | | |
|---|---|---|---|---|---|---|---|---|---|---|---|---|---|
| | n (female %) | | | Observed BMI (kg/$m^2$) | | | | ΔBMI (kg/$m^2$) partial interaction | | | | | |
| | Bottom PRS decile | Top PRS decile | | Bottom PRS decile | Top PRS decile | Contrast | p-value | | Contrast | p-value | | Bottom PRS decile | Top PRS decile |
| 1975 | 646 (55) | 645 (57) | 1975 | 22.1 (0.1) | 24.6 (0.1) | 2.5 (0.2) | <0.001 | 1975 vs 1981 | 0.3 (0.3) | 0.26 | PRS, z-score | -1.7 (0.4) | 1.8 (0.4) |
| 1981 | 612 (56) | 602 (58) | 1981 | 22.7 (0.1) | 25.6 (0.2) | 2.9 (0.2) | <0.001 | 1981 vs 1990 | 0.4 (0.3) | 0.18 | Expected BMI 1975, kg/$m^2$ | 21.8 (1.6) | 24.5 (1.6) |
| 1990 | 496 (56) | 473 (58) | 1990 | 23.7 (0.2) | 27.0 (0.2) | 3.3 (0.2) | <0.001 | 1990 vs 2011 | 1.4 (0.4) | 0.0012 | | | |
| 2011 | 229 (57) | 237 (56) | 2011 | 24.8 (0.2) | 29.4 (0.2) | 4.7 (0.3) | <0.001 | | | | | | |

Values are frequency (percentages) or mean (Delta-method standard error [SE]) and p-values were derived from contrast tests and post-hoc tests of partial interaction from the repeated measures mixed-effects linear regressions. PRS = polygenic risk score, n = number of individuals, BMI = body mass index, ΔBMI = BMI difference.

Table S2b: Adjusted values of characteristics of twin individuals in bottom and top PRS deciles

| Comparison of characteristics between twins in top and bottom PRS decile | | | | | | | |
|---|---|---|---|---|---|---|---|
| | Observed BMI (kg/$m^2$) | | | | ΔBMI (kg/$m^2$) partial interaction | | |
| | Bottom PRS decile | Top PRS decile | Contrast | p-value | | Contrast | p-value |
| 1975 | 22.1 (0.1) | 24.6 (0.1) | 2.5 (0.2) | <0.001 | 1975 vs 1981 | 0.4 (0.1) | <0.001 |
| 1981 | 22.7 (0.2) | 25.6 (0.1) | 2.9 (0.2) | <0.001 | 1981 vs 1990 | 0.5 (0.2) | 0.0035 |
| 1990 | 23.8 (0.2) | 27.2 (0.2) | 3.4 (0.3) | <0.001 | 1990 vs 2011 | 0.9 (0.3) | 0.0017 |
| 2011 | 25.7 (0.3) | 29.9 (0.3) | 4.3 (0.4) | <0.001 | | | |

Values are mean (Delta-method std. err.) and p-values were derived from contrast tests and post-hoc tests of partial interaction from the repeated measures mixed-effects linear regressions. PRS = polygenic risk score, n = number of individuals, BMI = body mass index, ΔBMI = BMI difference.

Table S3a: Unadjusted means of observed Body mass index values by polygenic risk score decile over time

| | **Body mass index (kg/m²)** | | | | |
|---|---|---|---|---|---|
| PRS percentile | 1975 | 1981 | 1990 | 2011 | ΔBMI 1975–2011, kg/m² |
| 0-10 | 22.1 (0.1) | 22.7 (0.1) | 23.7 (0.2) | 24.8 (0.2) | 2.7 [2.5, 2.9] |
| 10-20 | 22.3 (0.1) | 23.1 (0.1) | 24.2 (0.2) | 24.8 (0.2) | 2.4 [2.2, 2.6] |
| 20-30 | 22.6 (0.1) | 23.2 (0.1) | 24.1 (0.2) | 25.4 (0.2) | 2.8 [2.6, 3.0] |
| 30-40 | 22.9 (0.1) | 23.6 (0.1) | 24.5 (0.2) | 26.1 (0.2) | 3.2 [3.0, 3.4] |
| 40-50 | 23.2 (0.1) | 23.9 (0.1) | 24.8 (0.2) | 25.9 (0.2) | 2.7 [2.5, 2.9] |
| 50-60 | 23.2 (0.1) | 23.9 (0.1) | 24.9 (0.2) | 26.5 (0.2) | 3.3 [3.1, 3.5] |
| 60-70 | 23.2 (0.1) | 23.9 (0.1) | 25.0 (0.2) | 26.4 (0.2) | 3.2 [3.0, 3.4] |
| 70-80 | 23.7 (0.1) | 24.4 (0.1) | 25.6 (0.2) | 27.4 (0.2) | 3.7 [3.5, 3.9] |
| 80-90 | 24.2 (0.1) | 25.0 (0.1) | 26.3 (0.2) | 28.2 (0.2) | 4.1 [3.9, 4.3] |
| 90-100 | 24.6 (0.1) | 25.6 (0.1) | 27.0 (0.2) | 29.4 (0.2) | 4.8 [4.7, 5.0] |

BMI values are mean (Delta-method std. err.) and ΔBMI values are mean [95% confidence interval].

Table S3b: Adjusted means body mass index values by polygenic risk score decile over time.

| | **Body mass index (kg/m²)** | | | | |
|---|---|---|---|---|---|
| PRS percentile | 1975 | 1981 | 1990 | 2011 | ΔBMI 1975–2011, kg/m² |
| 0-10 | 22.1 (0.1) | 22.7 (0.1) | 23.8 (0.2) | 25.7 (0.2) | 3.6 [3.4, 3.8] |
| 10-20 | 22.4 (0.1) | 23.2 (0.1) | 24.4 (0.2) | 26.0 (0.2) | 3.6 [3.4, 3.9] |
| 20-30 | 22.7 (0.1) | 23.3 (0.1) | 24.4 (0.2) | 26.6 (0.2) | 3.9 [3.7, 4.2] |
| 30-40 | 22.9 (0.1) | 23.7 (0.1) | 24.8 (0.2) | 27.2 (0.2) | 4.3 [4.1, 4.5] |
| 40-50 | 23.1 (0.1) | 23.8 (0.1) | 25.0 (0.2) | 27.1 (0.2) | 4.0 [3.7, 4.2] |
| 50-60 | 23.1 (0.1) | 23.9 (0.1) | 25.1 (0.2) | 27.6 (0.2) | 4.4 [4.2, 4.7] |
| 60-70 | 23.3 (0.1) | 24.0 (0.1) | 25.3 (0.2) | 27.5 (0.2) | 4.2 [3.9, 4.4] |
| 70-80 | 23.6 (0.1) | 24.5 (0.1) | 25.8 (0.2) | 28.4 (0.2) | 4.8 [4.5, 5.0] |
| 80-90 | 24.2 (0.1) | 25.0 (0.1) | 26.4 (0.2) | 29.2 (0.2) | 5.0 [4.8, 5.3] |
| 90-100 | 24.6 (0.1) | 25.6 (0.1) | 27.1 (0.2) | 30.0 (0.2) | 5.4 [5.2, 5.6] |

BMI values are mean (Delta-method std. err.) and ΔBMI values are mean [95% confidence interval].

Table S3c: Number of individuals by polygenic risk score decile over time.

| PRS percentile | 1975 | 1981 | 1990 | 2011 |
|---|---|---|---|---|
| 0-10 | 646 | 612 | 496 | 229 |
| 10-20 | 646 | 604 | 479 | 211 |
| 20-30 | 645 | 616 | 484 | 206 |
| 30-40 | 646 | 612 | 489 | 218 |
| 40-50 | 644 | 605 | 464 | 196 |
| 50-60 | 647 | 616 | 462 | 197 |
| 60-70 | 645 | 606 | 473 | 236 |
| 70-80 | 645 | 608 | 487 | 212 |
| 80-90 | 645 | 596 | 483 | 212 |
| 90-100 | 645 | 602 | 473 | 237 |

Values are frequency.

Table S4a: Regression analyses between $PRS_{BMI}$ and BMI in 8,309 MZ and DZ twin individuals

| | β BMI ~ $PRS_{BMI}$ | $R^2$ | P-value | PRS min | PRS max | PRS range | BMI over PRS range |
|---|---|---|---|---|---|---|---|
| $PRS_{BMI}$ (z-score [95% CI]) | 0.71 [0.64; 0.78] | 0.049 | <0.001 | -3.48 | 3.68 | 7.16 | 5.1 [4.6; 5.7] |
| Intercept | 23.3 [23.3; 23.4] | | | | | | |

Beta (β) and [95% confidence interval, 95% CI], BMI = body mass index, $PRS_{BMI}$ = polygenic risk score for body mass index, min = minimum, max = maximum

Table S4b: Regression analyses between $PRS_{BMI}$ and BMI of 8,309 MZ and DZ twin individuals, corrected for age and sex

| | β BMI | $R^2$ | P-value | Min | Max | Range | BMI over range |
|---|---|---|---|---|---|---|---|
| $PRS_{BMI}$ (z-score, [95% CI]) | 0.78 [0.73; 0.84] | 0.293 | <0.001 | -3.48 | 3.68 | 7.16 | 5.6 [5.2; 6.0] |
| Age, years | 0.15 [0.14; 0.15] | | <0.001 | 18 | 75 | 57 | 8.4 |
| Sex | -1.53 [-1.64; -1.41] | | <0.001 | 1 (men) | 2 (women) | 1 | -1.5 |
| Intercept | 20.6 [20.4; 20.9] | | <0.001 | | | | |

Beta (β) and [95% confidence interval, 95% CI], BMI = body mass index, $PRS_{BMI}$ = polygenic risk score for body mass index, min = minimum, max = maximum

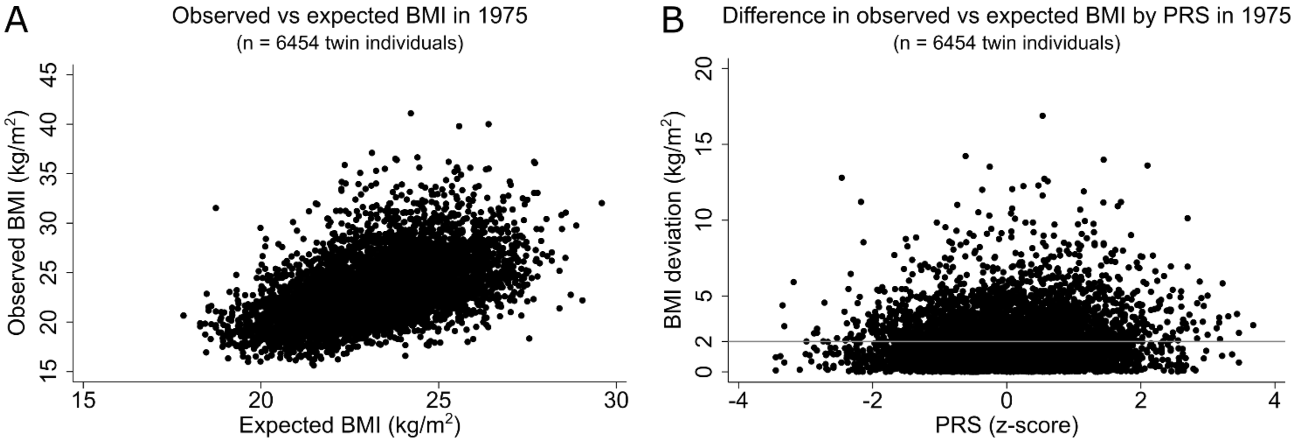

Figure S1: Scatterplots of observed body mass index (BMI) against expected BMI (A) and the absolute difference between observed and expected BMI based on polygenic risk scores (PRS) plotted against PRS (B) in all twin individuals from complete twin pairs in 1975. The gray line at 2.0 kg/m² represents the average BMI deviation.

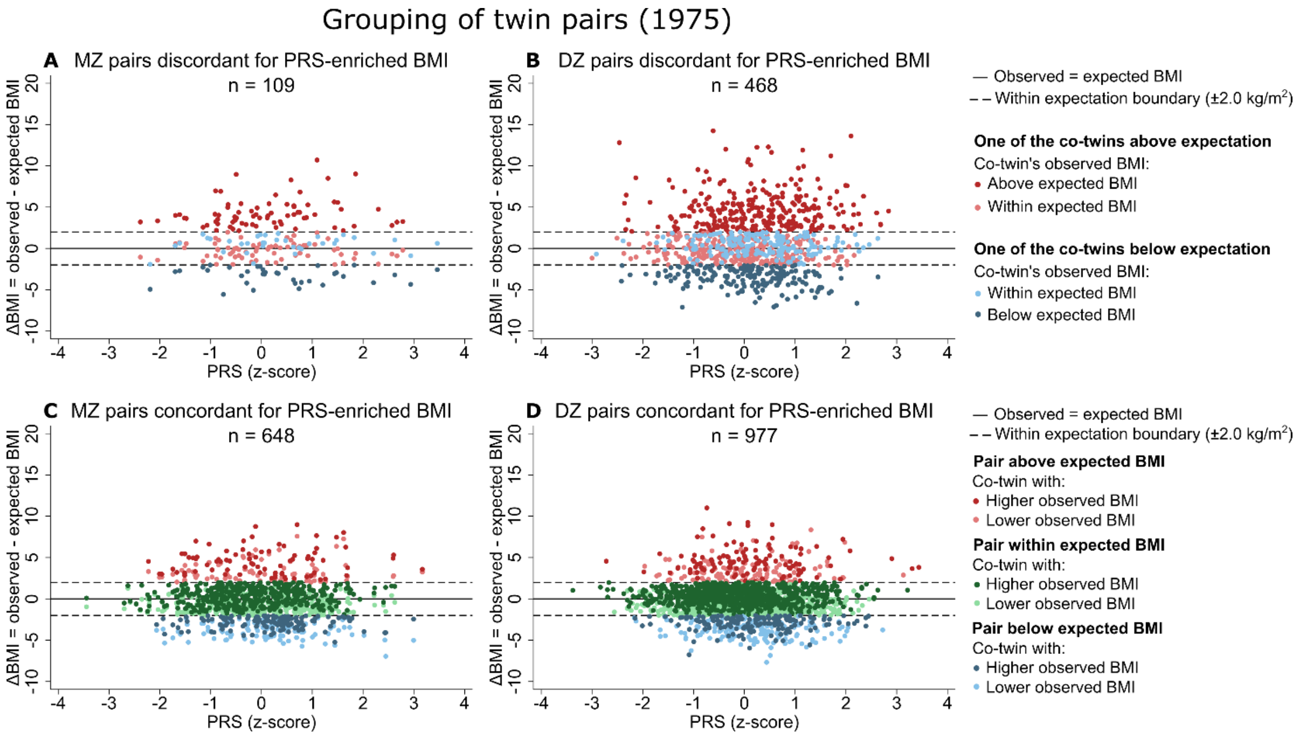

Figure S2: Categorizing the BMI difference (ΔBMI) between the observed body mass index (BMI) and the expected BMI in monozygotic (MZ) and dizygotic (DZ) twin pairs discordant and concordant for PRS-enriched BMI (based on observed and expected BMI values). n = number of pairs.

Table S5: Number of twin pairs that participated per timepoint in subgroups of monozygotic and dizygotic twin pairs discordant for PRS-enriched BMI

| | **Twin pairs discordant for PRS-enriched BMI** | | | |
|---|---|---|---|---|
| | MZ twin pairs with one of the co-twin's observed BMI | | DZ twin pairs with one of the co-twin's observed BMI | |
| | Below expectation | Above expectation | Below expectation | Above expectation |
| 1975, n (female % of n) | 36 (64) | 73 (53) | 179 (57) | 289 (59) |
| 1981, n (female % of n) | 33 (64) | 69 (54) | 159 (53) | 249 (61) |
| 1990, n (female % of n) | 20 (60) | 49 (51) | 103 (56) | 182 (61) |
| 2011, n (female % of n) | 11 (64) | 23 (52) | 26 (69) | 57 (51) |

Values are frequency (percentage). BMI = body mass index, PRS = polygenic risk score, discordant for PRS-enriched BMI = large within-twin-pair differences in body mass index (≥ 3 $kg/m^2$) considering both observed and expected BMI, n = number of pairs, MZ = monozygotic, DZ = dizygotic.

Table S6: Number of twin pairs that participated per timepoint in subgroups of monozygotic and dizygotic twin pairs concordant for PRS-enriched BMI

| | **Twin pairs concordant for PRS-enriched BMI** | | | | | |
|---|---|---|---|---|---|---|
| | MZ twin pairs' observed BMI in both co-twins | | | DZ twin pairs' observed BMI in both co-twins | | |
| | Below expectation | Within expectation | Above expectation | Below expectation | Within expectation | Above expectation |
| 1975, n (female %) | 145 (66) | 463 (54) | 88 (63) | 162 (53) | 702 (52) | 113 (52) |
| 1981, n (female %) | 139 (68) | 433 (55) | 80 (64) | 149 (55) | 640 (54) | 105 (52) |
| 1990, n (female %) | 102 (69) | 337 (56) | 60 (62) | 100 (50) | 448 (54) | 71 (58) |
| 2011, n (female %) | 48 (63) | 210 (58) | 37 (73) | 32 (50) | 209 (54) | 24 (50) |

Values are frequency (percentage). BMI = body mass index, PRS = polygenic risk score, concordant for PRS-enriched BMI = small within-twin-pair differences in body mass index (< 3 $kg/m^2$) considering both observed and expected BMI, n = number of pairs, MZ = monozygotic, DZ = dizygotic.

Table S7: Polygenic risk score and expected BMI in 1975 in subgroups of MZ and DZ twin pairs discordant for PRS-enriched BMI

| | **Twin pairs discordant for PRS-enriched BMI** | | | | | | | |
|---|---|---|---|---|---|---|---|---|
| | MZ twin pairs with one of the co-twin's observed BMI | | DZ twin pairs with one of the co-twin's observed BMI | | | | | |
| | Below expectation (n = 36) | Above expectation (n = 73) | Below expectation (n = 179) | | | Above expectation (n = 289) | | |
| | Both co-twins below and within expectation | Both co-twins above and within expectation | Co-twin below expectation | Co-twin within expectation | p-value | Co-twin within expectation | Co-twin above expectation | p-value |
| PRS, z-score | 0.4 (1.3) | 0.1 (1.1) | 0.1 (0.9) | 0.3 (0.9) | <0.001 | -0.1 (1.0) | 0.3 (1.0) | <0.001 |
| Expected BMI, $kg/m^2$ | 23.5 (1.7) | 23.5 (1.6) | 23.8 (1.8) | 24.0 (1.8) | 0.31 | 23.3 (1.6) | 23.6 (1.7) | 0.022 |

Values are mean (SD) and t-tests gave p-values. BMI = body mass index, PRS = polygenic risk score, discordant for PRS-enriched BMI = large within-twin-pair differences in body mass index (≥ 3 $kg/m^2$) considering both observed and expected BMI, MZ = monozygotic, DZ = dizygotic, n = number of pairs, ΔBMI = observed BMI minus expected BMI, |ΔBMI| = absolute values of observed BMI minus expected BMI

Table S8: Polygenic risk scores and expected BMI in 1975 in subgroups of MZ twin pairs concordant for PRS-enriched BMI

| | **MZ twin pairs concordant for PRS-enriched BMI** | | |
|---|---|---|---|
| | Both co-twins in MZ pairs below expectation (n = 136) | Both co-twins in MZ pairs within expectation (n = 432) | Both co-twins in MZ pairs above expectation (n = 80) |
| PRS, z-score | 0.1 (0.9) | -0.2 (1.0) | 0.0 (1.1) |
| Expected BMI, $kg/m^2$ | 23.1 (1.7) | 22.6 (1.7) | 22.9 (1.7) |

Values are mean (SD) and t-tests gave p-values. MZ = monozygotic, BMI = body mass index, PRS = polygenic risk score, concordant for PRS-enriched BMI = small within-twin-pair differences in body mass index (< 3 $kg/m^2$) considering both observed and expected BMI, n = number of pairs

Table S9: Polygenic risk scores and expected BMI in 1975 in subgroups of DZ twin pairs concordant for PRS-enriched BMI

| | **DZ twin pairs concordant for PRS-enriched BMI** | | | | | | | | |
|---|---|---|---|---|---|---|---|---|---|
| | DZ pair below expectation (n = 162) | | | DZ pair within expectation (n = 702) | | | DZ pair above expectation (n = 113) | | |
| | Co-twin with lower BMI | Co-twin with higher BMI | p-value | Co-twin with lower BMI | Co-twin with higher BMI | p-value | Co-twin with lower BMI | Co-twin with higher BMI | p-value |
| PRS, z-score | 0.2 (0.9) | 0.2 (0.9) | 0.81 | -0.1 (1.0) | -0.1 (0.9) | 0.016 | 0.1 (1.0) | 0.1 (0.9) | 0.82 |
| Expected BMI, $kg/m^2$ | 23.7 (1.7) | 23.7 (1.7) | 0.96 | 23.0 (1.8) | 23.1 (1.7) | 0.59 | 23.6 (1.7) | 23.6 (1.7) | 0.96 |

Values are mean (SD) and t-tests gave p-values. DZ = dizygotic, BMI = body mass index, PRS = polygenic risk score, concordant for PRS-enriched BMI = small within-twin-pair differences in body mass index (< 3 $kg/m^2$) considering both observed and expected BMI, n = number of pairs.

Table S10a: Unadjusted values from contrast tests that compared observed body mass index at each timepoint within subgroups of monozygotic twin pairs discordant for PRS-enriched BMI

**MZ twin pairs discordant for PRS-enriched BMI**

| | One of the MZ co-twin's observed BMI | | | | | | | |
|---|---|---|---|---|---|---|---|---|
| | Below expectation (n = 36) | | | | Above expectation (n = 73) | | | |
| | Co-twin below expectation | Co-twin within expectation | Contrast | p-value | Co-twin within expectation | Co-twin above expectation | Contrast | p-value |
| 1975, kg/m$^2$ | 20.2 (0.4) | 24.1 (0.4) | 3.8 (0.6) | <0.001 | 23.3 (0.4) | 27.8 (0.4) | 4.5 (0.5) | <0.001 |
| 1981, kg/m$^2$ | 21.2 (0.4) | 24.0 (0.5) | 2.7 (0.6) | <0.001 | 24.7 (0.4) | 27.3 (0.4) | 2.5 (0.5) | <0.001 |
| 1990, kg/m$^2$ | 21.3 (0.6) | 24.6 (0.5) | 3.3 (0.8) | <0.001 | 26.5 (0.4) | 28.4 (0.4) | 1.9 (0.6) | 0.002 |
| 2011, kg/m$^2$ | 23.3 (0.8) | 26.4 (0.8) | 3.1 (1.1) | 0.006 | 28.2 (0.6) | 31.3 (0.6) | 3.1 (0.9) | 0.001 |
| ΔBMI 1975–2011, kg/m$^2$ | 3.0 [2.4, 3.7] | 2.3 [1.6, 3.0] | | | 4.9 [4.4, 5.4] | 3.5 [3.0, 4.0] | | |
| ΔBMI 1975–2011, p-value | 0.001 | 0.011 | | | <0.001 | <0.001 | | |

BMI values are mean (Delta-method std. err.), ΔBMI values are mean [95% confidence interval], and contrast tests gave mean (SE) values and p-values. MZ = monozygotic, BMI = body mass index, PRS = polygenic risk score, discordant for PRS-enriched BMI = large within-twin-pair differences in body mass index (≥ 3 kg/m$^2$) considering both observed and expected BMI, n = number of pairs, ΔBMI = $BMI_{2011}$ – $BMI_{1975}$.

Table S10b: Adjusted values from contrast tests that compared observed body mass index at each timepoint within subgroups of monozygotic twin pairs discordant for PRS-enriched BMI

**MZ twin pairs discordant for PRS-enriched BMI**

| | One of the MZ co-twin's observed BMI | | | | | | | |
|---|---|---|---|---|---|---|---|---|
| | Below expectation (n = 36) | | | | Above expectation (n = 73) | | | |
| | Co-twin below expectation | Co-twin within expectation | Contrast | p-value | Co-twin within expectation | Co-twin above expectation | Contrast | p-value |
| 1975, kg/m$^2$ | 20.2 (0.3) | 24.1 (0.3) | 3.8 (0.1) | <0.001 | 23.3 (0.2) | 27.8 (0.2) | 4.5 (0.2) | <0.001 |
| 1981, kg/m$^2$ | 21.3 (0.5) | 24.0 (0.5) | 2.7 (0.5) | <0.001 | 24.8 (0.3) | 27.3 (0.3) | 2.5 (0.3) | <0.001 |
| 1990, kg/m$^2$ | 21.9 (0.6) | 25.2 (0.5) | 3.3 (0.6) | <0.001 | 26.5 (0.5) | 28.6 (0.5) | 2.1 (0.6) | 0.001 |
| 2011, kg/m$^2$ | 24.6 (1.0) | 27.8 (1.0) | 3.2 (1.1) | 0.003 | 28.7 (0.7) | 32.0 (0.7) | 3.2 (1.0) | 0.001 |
| ΔBMI 1975–2011, kg/m$^2$ | 4.3 [3.1, 5.6] | 3.7 [2.5, 5.0] | | | 5.5 [4.5, 6.4] | 4.2 [3.2, 5.2] | | |
| ΔBMI 1975–2011, p-value | <0.001 | <0.001 | | | <0.001 | <0.001 | | |

BMI values are mean (Delta-method std. err.), ΔBMI values are mean [95% confidence interval], and contrast tests gave mean (SE) values and p-values. MZ = monozygotic, discordant for PRS-enriched BMI = large within-twin-pair differences in body mass index (≥ 3 kg/m$^2$) considering both observed and expected BMI, BMI = body mass index, PRS = polygenic risk score, n = number of pairs, ΔBMI = $BMI_{2011}$ – $BMI_{1975}$.

Table S11a: Unadjusted values from contrast tests that compared observed body mass index at each timepoint within subgroups of dizygotic twin pairs discordant for PRS-enriched BMI

| | DZ twin pairs discordant for PRS-enriched BMI | | | | | | | |
|---|---|---|---|---|---|---|---|---|
| | One of the DZ co-twin's observed BMI | | | | | | | |
| | Below expectation (n = 179) | | | | Above expectation (n = 289) | | | |
| | Co-twin below expectation | Co-twin within expectation | Contrast | p-value | Co-twin within expectation | Co-twin above expectation | Contrast | p-value |
| 1975, kg/m$^2$ | 20.2 (0.2) | 24.4 (0.2) | 4.2 (0.3) | <0.001 | 23.0 (0.2) | 28.4 (0.2) | 5.3 (0.3) | <0.001 |
| 1981, kg/m$^2$ | 21.2 (0.2) | 25.0 (0.2) | 3.8 (0.3) | <0.001 | 24.0 (0.2) | 28.9 (0.2) | 4.9 (0.3) | <0.001 |
| 1990, kg/m$^2$ | 21.9 (0.2) | 26.0 (0.2) | 4.1 (0.3) | <0.001 | 25.0 (0.2) | 30.1 (0.2) | 5.2 (0.3) | <0.001 |
| 2011, kg/m$^2$ | 23.8 (0.4) | 27.8 (0.4) | 4.0 (0.6) | <0.001 | 26.6 (0.4) | 32.0 (0.4) | 5.4 (0.5) | <0.001 |
| ΔBMI 1975–2011, kg/m$^2$ | 3.5 [3.0, 4.0] | 3.4 [2.9, 3.9] | | | 3.6 [3.2, 3.9] | 3.6 [3.2, 4.0] | | |
| ΔBMI 1975–2011, p-value | <0.001 | <0.001 | | | <0.001 | <0.001 | | |

BMI values are mean (Delta-method std. err.), ΔBMI values are mean [95% confidence interval], and contrast tests gave mean (SE) values and p-values. DZ = dizygotic, discordant for PRS-enriched BMI = large within-twin-pair differences in body mass index (≥ 3 kg/m$^2$) considering both observed and expected BMI, BMI = body mass index, PRS = polygenic risk score, n = number of pairs, ΔBMI = $BMI_{2011}$ – $BMI_{1975}$.

Table S11b: Adjusted values from contrast tests that compared observed body mass index at each timepoint within subgroups of dizygotic twin pairs discordant for PRS-enriched BMI

| | DZ twin pairs discordant for PRS-enriched BMI | | | | | | | |
|---|---|---|---|---|---|---|---|---|
| | One of the DZ co-twin's observed BMI | | | | | | | |
| | Below expectation (n = 179) | | | | Above expectation (n = 289) | | | |
| | Co-twin below expectation | Co-twin within expectation | Contrast | p-value | Co-twin within expectation | Co-twin above expectation | Contrast | p-value |
| 1975, kg/m$^2$ | 20.2 (0.1) | 24.4 (0.1) | 4.2 (0.1) | <0.001 | 23.0 (0.1) | 28.4 (0.1) | 5.3 (0.1) | <0.001 |
| 1981, kg/m$^2$ | 21.2 (0.2) | 24.9 (0.2) | 3.7 (0.2) | <0.001 | 24.0 (0.2) | 28.8 (0.2) | 4.8 (0.2) | <0.001 |
| 1990, kg/m$^2$ | 22.4 (0.3) | 26.3 (0.3) | 4.0 (0.4) | <0.001 | 25.3 (0.2) | 30.3 (0.2) | 5.0 (0.3) | <0.001 |
| 2011, kg/m$^2$ | 25.7 (0.6) | 29.2 (0.6) | 3.5 (0.8) | <0.001 | 27.4 (0.4) | 32.6 (0.4) | 5.2 (0.6) | <0.001 |
| ΔBMI 1975–2011, kg/m$^2$ | 5.5 [4.6, 6.3] | 4.8 [4.0, 5.6] | | | 4.3 [3.8, 4.9] | 4.2 [3.6, 4.8] | | |
| ΔBMI 1975–2011, p-value | <0.001 | <0.001 | | | <0.001 | <0.001 | | |

BMI values are mean (Delta-method std. err.), ΔBMI values are mean [95% confidence interval], and contrast tests gave mean (SE) values and p-values. DZ = dizygotic, discordant for PRS-enriched BMI = large within-twin-pair differences in body mass index (≥ 3 kg/m$^2$) considering both observed and expected BMI, BMI = body mass index, PRS = polygenic risk score, n = number of pairs, ΔBMI = $BMI_{2011}$ – $BMI_{1975}$.

Table S12a: Unadjusted values from post-hoc tests of partial interaction between co-twin subgroup and BMI between adjacent timepoints in monozygotic and dizygotic twin pairs discordant for PRS-enriched BMI

| | **BMI differences between co-twins and adjacent timepoints in twin pairs discordant for PRS-enriched BMI** | | | | | | | |
|---|---|---|---|---|---|---|---|---|
| | MZ co-twins below vs within (n = 36) | | MZ co-twins above vs within (n = 73) | | DZ co-twins below vs within (n = 179) | | DZ co-twins above vs within (n = 289) | |
| | Contrast | p-value | Contrast | p-value | Contrast | p-value | Contrast | p-value |
| 1975 vs 1981, kg/m$^2$ | -1.1 (0.9) | 0.21 | -2.0 (0.8) | 0.009 | -0.3 (0.4) | 0.39 | -0.4 (0.4) | 0.25 |
| 1981 vs 1990, kg/m$^2$ | 0.6 (1.0) | 0.56 | -0.6 (0.8) | 0.44 | 0.3 (0.4) | 0.45 | 0.3 (0.4) | 0.49 |
| 1990 vs 2011, kg/m$^2$ | -0.2 (1.3) | 0.87 | 1.2 (1.1) | 0.27 | -0.1 (0.7) | 0.87 | 0.2 (0.6) | 0.76 |

Values are mean (SE) and post-hoc tests of partial interaction gave p-values. Discordant for PRS-enriched BMI = large within-twin-pair differences in body mass index (≥ 3 kg/m$^2$) considering both observed and expected BMI, BMI = body mass index, PRS = polygenic risk score, n = number of pairs, MZ = monozygotic, DZ = dizygotic.

Table S12b: Adjusted values from post-hoc tests of partial interaction between co-twin subgroup and BMI between adjacent timepoints in monozygotic and dizygotic twin pairs discordant for PRS-enriched BMI

| | **BMI differences between co-twins and adjacent timepoints in twin pairs discordant for PRS-enriched BMI** | | | | | | | |
|---|---|---|---|---|---|---|---|---|
| | MZ co-twins below vs within (n = 36) | | MZ co-twins above vs within (n = 73) | | DZ co-twins below vs within (n = 179) | | DZ co-twins above vs within (n = 289) | |
| | Contrast | p-value | Contrast | p-value | Contrast | p-value | Contrast | p-value |
| 1975 vs 1981, kg/m$^2$ | -1.1 (0.5) | 0.015 | -2.0 (0.3) | <0.001 | -0.5 (0.2) | 0.004 | -0.5 (0.2) | 0.007 |
| 1981 vs 1990, kg/m$^2$ | 0.6 (0.5) | 0.24 | -0.5 (0.5) | 0.37 | 0.3 (0.4) | 0.43 | 0.2 (0.2) | 0.48 |
| 1990 vs 2011, kg/m$^2$ | -0.1 (1.2) | 0.95 | 1.2 (0.9) | 0.19 | -0.5 (0.8) | 0.55 | 0.2 (0.5) | 0.69 |

Values are mean (SE) and post-hoc tests of partial interaction gave p-values. Discordant for PRS-enriched BMI = large within-twin-pair differences in body mass index (≥ 3 kg/m$^2$) considering both observed and expected BMI, BMI = body mass index, PRS = polygenic risk score, n = number of pairs, MZ = monozygotic, DZ = dizygotic.

Table S13a: Unadjusted values from contrast tests that compared observed body mass index at each timepoint within subgroups of monozygotic twin pairs concordant for PRS-enriched BMI

**Observed BMI (kg/m²) in MZ twin pairs concordant for PRS-enriched BMI**

| | MZ pair below expectation (n = 136) | | | | MZ pair within expectation (n = 432) | | | | MZ pair above expectation (n = 80) | | | |
|---|---|---|---|---|---|---|---|---|---|---|---|---|
| | Co-twin with lower BMI | Co-twin with higher BMI | Contrast | p-value | Co-twin with lower BMI | Co-twin with higher BMI | Contrast | p-value | Co-twin with lower BMI | Co-twin with higher BMI | Contrast | p-value |
| 1975 | 19.4 (0.2) | 20.2 (0.2) | 0.9 (0.3) | 0.001 | 22.0 (0.1) | 22.9 (0.1) | 0.9 (0.2) | <0.001 | 26.3 (0.4) | 27.5 (0.4) | 1.2 (0.6) | 0.028 |
| 1981 | 20.6 (0.2) | 20.8 (0.2) | 0.2 (0.3) | 0.37 | 23.0 (0.1) | 23.7 (0.1) | 0.7 (0.2) | <0.001 | 27.2 (0.4) | 27.7 (0.4) | 0.5 (0.6) | 0.35 |
| 1990 | 21.5 (0.2) | 21.6 (0.2) | 0.04 (0.3) | 0.88 | 24.0 (0.1) | 24.8 (0.1) | 0.7 (0.2) | <0.001 | 28.6 (0.4) | 29.3 (0.4) | 0.7 (0.6) | 0.27 |
| 2011 | 23.3 (0.3) | 23.6 (0.3) | 0.2 (0.4) | 0.59 | 25.9 (0.2) | 26.2 (0.2) | 0.3 (0.3) | 0.18 | 29.0 (0.6) | 30.6 (0.6) | 1.6 (0.8) | 0.062 |
| ΔBMI | 4.0 [3.7, 4.2] | 3.3 [3.1, 3.6] | | | 3.9 [3.8, 4.0] | 3.3 [3.2, 3.4] | | | 2.7 [2.3, 3.1] | 3.1 [2.7, 3.5] | | |
| ΔBMI *P* | <0.001 | <0.001 | | | <0.001 | <0.001 | | | <0.001 | <0.001 | | |

BMI values are mean (Delta-method std. err.), ΔBMI values are mean [95% confidence interval], and contrast tests gave mean (SE) values and p-values. BMI = body mass index, MZ = monozygotic, concordant for PRS-enriched BMI = small within-twin-pair differences in body mass index (< 3 kg/m²) considering both observed and expected BMI, PRS = polygenic risk score, n = number of pairs, ΔBMI = $BMI_{2011}$ – $BMI_{1975}$, ΔBMI *P* = p-values for $BMI_{2011}$ – $BMI_{1975}$ by co-twin group.

Table S13b: Adjusted values from contrast tests that compared observed body mass index at each timepoint within subgroups of monozygotic twin pairs concordant for PRS-enriched BMI

**Observed BMI (kg/m²) in MZ twin pairs concordant for PRS-enriched BMI**

| | MZ pair below expectation (n = 136) | | | | MZ pair within expectation (n = 432) | | | | MZ pair above expectation (n = 80) | | | |
|---|---|---|---|---|---|---|---|---|---|---|---|---|
| | Co-twin with lower BMI | Co-twin with higher BMI | Contrast | p-value | Co-twin with lower BMI | Co-twin with higher BMI | Contrast | p-value | Co-twin with lower BMI | Co-twin with higher BMI | Contrast | p-value |
| 1975 | 19.4 (0.1) | 20.2 (0.1) | 0.9 (0.1) | <0.001 | 22.0 (0.1) | 22.9 (0.1) | 0.9 (0.04) | <0.001 | 26.3 (0.3) | 27.5 (0.3) | 1.2 (0.1) | <0.001 |
| 1981 | 20.6 (0.2) | 20.8 (0.2) | 0.2 (0.1) | 0.19 | 23.0 (0.1) | 23.7 (0.1) | 0.7 (0.1) | <0.001 | 27.2 (0.3) | 27.7 (0.3) | 0.4 (0.3) | 0.22 |
| 1990 | 21.8 (0.2) | 21.8 (0.2) | -0.03 (0.2) | 0.88 | 24.1 (0.1) | 24.8 (0.1) | 0.7 (0.1) | <0.001 | 28.6 (0.4) | 29.2 (0.4) | 0.5 (0.5) | 0.30 |
| 2011 | 23.6 (0.4) | 24.4 (0.4) | 0.8 (0.6) | 0.13 | 26.5 (0.2) | 26.9 (0.2) | 0.5 (0.3) | 0.11 | 30.5 (0.7) | 31.8 (0.7) | 1.4 (0.9) | 0.11 |
| ΔBMI | 4.2 [3.7, 4.7] | 4.2 [3.6, 4.7] | | | 4.4 [4.2, 4.7] | 4.0 [3.7, 4.3] | | | 4.2 [3.4, 5.0] | 4.4 [3.6, 5.1] | | |
| ΔBMI *P* | <0.001 | <0.001 | | | <0.001 | <0.001 | | | <0.001 | <0.001 | | |

BMI values are mean (Delta-method std. err.), ΔBMI values are mean [95% confidence interval], and contrast tests gave mean (SE) values and p-values. BMI = body mass index, MZ = monozygotic, concordant for PRS-enriched BMI = small within-twin-pair differences in body mass index (< 3 kg/m²) considering both observed and expected BMI, PRS = polygenic risk score, n = number of pairs, ΔBMI = $BMI_{2011}$ – $BMI_{1975}$, ΔBMI *P* = p-values for $BMI_{2011}$ – $BMI_{1975}$ by co-twin group.

Table S14a: Unadjusted values from contrast tests that compared observed body mass index at each timepoint within subgroups of dizygotic twin pairs concordant for PRS-enriched BMI

**Observed BMI (kg/m²) in DZ twin pairs concordant for PRS-enriched BMI**

| | DZ pair below expectation (n = 162) | | | | DZ pair within expectation (n = 702) | | | | DZ pair above expectation (n = 113) | | | |
|---|---|---|---|---|---|---|---|---|---|---|---|---|
| | Co-twin with lower BMI | Co-twin with higher BMI | Contrast | p-value | Co-twin with lower BMI | Co-twin with higher BMI | Contrast | p-value | Co-twin with lower BMI | Co-twin with higher BMI | Contrast | p-value |
| 1975 | 19.9 (0.2) | 20.8 (0.2) | 0.9 (0.2) | <0.001 | 22.4 (0.1) | 23.4 (0.1) | 1.0 (0.1) | <0.001 | 27.0 (0.3) | 28.3 (0.3) | 1.2 (0.4) | 0.002 |
| 1981 | 20.6 (0.2) | 21.6 (0.2) | 0.9 (0.2) | <0.001 | 23.2 (0.1) | 24.0 (0.1) | 0.7 (0.1) | <0.001 | 27.9 (0.3) | 28.6 (0.3) | 0.7 (0.4) | 0.10 |
| 1990 | 21.8 (0.2) | 22.6 (0.2) | 0.7 (0.3) | 0.007 | 24.3 (0.1) | 25.0 (0.1) | 0.7 (0.2) | <0.001 | 28.8 (0.3) | 29.6 (0.3) | 0.8 (0.5) | 0.071 |
| 2011 | 23.8 (0.3) | 24.5 (0.3) | 0.7 (0.5) | 0.12 | 25.9 (0.2) | 26.5 (0.2) | 0.6 (0.2) | 0.009 | 30.7 (0.6) | 31.2 (0.6) | 0.5 (0.8) | 0.56 |
| ΔBMI | 3.9 [3.6, 4.2] | 3.7 [3.4, 4.0] | | | 3.5 [3.3, 3.6] | 3.1 [2.9, 3.2] | | | 3.7 [3.1, 4.2] | 2.9 [2.3, 3.4] | | |
| ΔBMI *P* | <0.001 | <0.001 | | | <0.001 | <0.001 | | | <0.001 | <0.001 | | |

BMI values are mean (Delta-method std. err.), ΔBMI values are mean [95% confidence interval], and contrast tests gave mean (SE) values and p-values. BMI = body mass index, DZ = dizygotic, concordant for PRS-enriched BMI = small within-twin-pair differences in body mass index (< 3 kg/m²) considering both observed and expected BMI, PRS = polygenic risk score, n = number of pairs, ΔBMI = $BMI_{2011}$ – $BMI_{1975}$, ΔBMI *P* = p-values for $BMI_{2011}$ – $BMI_{1975}$ by co-twin group.

Table S14b: Adjusted values from contrast tests that compared observed body mass index at each timepoint within subgroups of dizygotic twin pairs concordant for PRS-enriched BMI

**Observed BMI (kg/m²) in DZ twin pairs concordant for PRS-enriched BMI**

| | DZ pair below expectation (n = 162) | | | | DZ pair within expectation (n = 702) | | | | DZ pair above expectation (n = 113) | | | |
|---|---|---|---|---|---|---|---|---|---|---|---|---|
| | Co-twin with lower BMI | Co-twin with higher BMI | Contrast | p-value | Co-twin with lower BMI | Co-twin with higher BMI | Contrast | p-value | Co-twin with lower BMI | Co-twin with higher BMI | Contrast | p-value |
| 1975 | 19.9 (0.1) | 20.8 (0.1) | 0.9 (0.1) | <0.001 | 22.4 (0.1) | 23.4 (0.1) | 1.0 (0.04) | <0.001 | 27.0 (0.2) | 28.3 (0.2) | 1.2 (0.1) | <0.001 |
| 1981 | 20.7 (0.2) | 21.6 (0.2) | 0.9 (0.1) | <0.001 | 23.3 (0.1) | 24.0 (0.1) | 0.7 (0.1) | <0.001 | 27.9 (0.3) | 28.6 (0.3) | 0.7 (0.3) | 0.014 |
| 1990 | 21.9 (0.2) | 22.9 (0.2) | 1.0 (0.2) | <0.001 | 24.5 (0.1) | 25.2 (0.1) | 0.7 (0.2) | <0.001 | 28.9 (0.4) | 30.1 (0.4) | 1.2 (0.4) | 0.005 |
| 2011 | 24.3 (0.4) | 25.4 (0.4) | 1.1 (0.6) | 0.066 | 26.8 (0.2) | 27.5 (0.2) | 0.7 (0.3) | 0.012 | 32.0 (0.7) | 32.7 (0.7) | 0.7 (1.0) | 0.45 |
| ΔBMI | 4.4 [3.8, 5.0] | 4.5 [4.0, 5.1] | | | 4.4 [4.1, 4.7] | 4.1 [3.9, 4.4] | | | 4.9 [4.0, 5.9] | 4.4 [3.4, 5.4] | | |
| ΔBMI *P* | <0.001 | <0.001 | | | <0.001 | <0.001 | | | <0.001 | <0.001 | | |

BMI values are mean (Delta-method std. err.), ΔBMI values are mean [95% confidence interval], and contrast tests gave mean (SE) values and p-values. BMI = body mass index, DZ = dizygotic, concordant for PRS-enriched BMI = small within-twin-pair differences in body mass index (< 3 kg/m²) considering both observed and expected BMI, PRS = polygenic risk score, n = number of pairs, ΔBMI = $BMI_{2011}$ – $BMI_{197}$, ΔBMI *P* = p-values for $BMI_{2011}$ – $BMI_{1975}$ by co-twin group.

Table S15a: Unadjusted values from post-hoc tests of partial interaction between co-twin subgroup and body mass index between adjacent timepoints in monozygotic and dizygotic twin pairs concordant for PRS-enriched BMI

| | **BMI differences between co-twins and adjacent timepoints in twin pairs concordant for PRS-enriched BMI** | | | | | | | | | | | |
|---|---|---|---|---|---|---|---|---|---|---|---|---|
| | MZ pair below (n = 136) | | MZ pair within (n = 432) | | MZ pair above (n = 80) | | DZ pair below (n = 162) | | DZ pair within (n = 702) | | DZ pair above (n = 113) | |
| | Contrast | p-value | Contrast | p-value | Contrast | p-value | Contrast | p-value | Contrast | p-value | Contrast | p-value |
| 1975 vs 1981, $kg/m^2$ | -0.6 (0.4) | 0.091 | -0.2 (0.3) | 0.42 | -0.7 (0.8) | 0.39 | 0.03 (0.3) | 0.94 | -0.3 (0.2) | 0.18 | -0.6 (0.6) | 0.31 |
| 1981 vs 1990, $kg/m^2$ | -0.2 (0.4) | 0.63 | -0.03 (0.3) | 0.92 | 0.2 (0.8) | 0.85 | -0.2 (0.4) | 0.62 | -0.001 (0.2) | 1.0 | 0.2 (0.6) | 0.79 |
| 1990 vs 2011, $kg/m^2$ | 0.2 (0.5) | 0.71 | -0.4 (0.3) | 0.23 | 0.9 (1.1) | 0.40 | -0.02 (0.5) | 0.98 | -0.1 (0.3) | 0.68 | -0.4 (0.9) | 0.68 |

Values are mean (SE) and post-hoc tests of partial interaction gave p-values. BMI = body mass index, concordant for PRS-enriched BMI = small within-twin-pair differences in body mass index (< 3 $kg/m^2$) considering both observed and expected BMI, PRS = polygenic risk score, MZ = monozygotic, n = number of pairs, DZ = dizygotic.

Table S15b: Adjusted values from post-hoc tests of partial interaction between co-twin subgroup and body mass index between adjacent timepoints in monozygotic and dizygotic twin pairs concordant for PRS-enriched BMI

| | **BMI differences between co-twins and adjacent timepoints in twin pairs concordant for PRS-enriched BMI** | | | | | | | | | | | |
|---|---|---|---|---|---|---|---|---|---|---|---|---|
| | MZ pair below (n = 136) | | MZ pair within (n = 432) | | MZ pair above (n = 80) | | DZ pair below (n = 162) | | DZ pair within (n = 702) | | DZ pair above (n = 113) | |
| | Contrast | p-value | Contrast | p-value | Contrast | p-value | Contrast | p-value | Contrast | p-value | Contrast | p-value |
| 1975 vs 1981, $kg/m^2$ | -0.7 (0.2) | <0.001 | -0.2 (0.1) | 0.037 | -0.8 (0.3) | 0.016 | 0.03 (0.1) | 0.80 | -0.3 (0.1) | 0.001 | -0.5 (0.3) | 0.060 |
| 1981 vs 1990, $kg/m^2$ | -0.2 (0.2) | 0.22 | -0.02 (0.1) | 0.87 | 0.1 (0.4) | 0.76 | 0.1 (0.2) | 0.74 | -0.01 (0.1) | 0.94 | 0.5 (0.4) | 0.20 |
| 1990 vs 2011, $kg/m^2$ | 0.9 (0.5) | 0.08 | -0.2 (0.3) | 0.41 | 0.8 (0.7) | 0.26 | 0.1 (0.5) | 0.89 | 0.01 (0.3) | 0.98 | -0.5 (1.0) | 0.61 |

Values are mean (SE) and post-hoc tests of partial interaction gave p-values. BMI = body mass index, concordant for PRS-enriched BMI = small within-twin-pair differences in body mass index (< 3 $kg/m^2$) considering both observed and expected BMI, PRS = polygenic risk score, MZ = monozygotic, n = number of pairs, DZ = dizygotic.
